# Analysis of factors associated early diagnosis in coronavirus disease 2019 (COVID-19)

**DOI:** 10.1101/2020.04.09.20059352

**Authors:** Jinwei Ai, Junyan Gong, Limin Xing, Renjiao He, Fangtao Tian, Juan Wang, Jun Wang, Shengduo Pei, Dongxuan Chen, Guoxin Huang, Meiling Zhang, Gaojing Qu, Wufeng Fan, Hongming Lin, Desheng Li, Bin Pei

**Affiliations:** Department of Evidence-Based Medicine Center, Xiangyang No.1 People’s Hospital, Hubei University of Medicine, Xiangyang 441000, China; The third ward of Orthopedic, Xiangyang No.1 People’s Hospital, Hubei University of Medicine, Xiangyang 441000, China; Department of endocrinology, Xiangyang No.1 People’s Hospital, Hubei University of Medicine, Xiangyang 441000, China; Department of Nursing, Xiangyang No.1 People’s Hospital, Hubei University of Medicine, Xiangyang 441000, China; Department of cardiology, Xiangyang No.1 People’s Hospital, Hubei University of Medicine, Xiangyang 441000, China; Department of Microbiology, Tumor and Cell biology, Karolinska Institute, Stockholm SE-17177, Sweden; Department of Statistics, Leiden University, Leiden 2333 CA, The Netherlands; Department of Medical Services Section, Xiangyang No.1 People’s Hospital, Hubei University of Medicine, Xiangyang 441000, China

**Keywords:** SARS-CoV-2, COVID-19, Logistic regression analysis, Diagnosis

## Abstract

**Background:** The pandemic of coronavirus disease 2019 (COVID-19) has become the first concern in international affairs as the novel coronavirus (SARS-CoV-2) is spreading all over the world at a terrific speed. The accuracy of early diagnosis is critical in the control of the spread of the virus. Although the real-time RT-PCR detection of the virus nucleic acid is the current golden diagnostic standard, it has high false negative rate when only apply single test.

**Objective:** Summarize the baseline characteristics and laboratory examination results of hospitalized COVID-19 patients. Analyze the factors that could interfere with the early diagnosis quantitatively to support the timely confirmation of the disease.

**Methods:** All suspected patients with COVID-19 were included in our study until Feb 9^th^, 2020. The last day of follow-up was Mar 20^th^, 2020. Throat swab real-time RT-PCR test was used to confirm SARS-CoV-2 infection. The difference between the epidemiological profile and first laboratory examination results of COVID-19 patients and non-COVID-19 patients were compared and analyzed by multiple logistic regression. Receiver operating characteristic (ROC) curve and area under curve (AUC) were used to assess the potential diagnostic value in factors, which had statistical differences in regression analysis.

**Results:** In total, 315 hospitalized patients were included. Among them, 108 were confirmed as COVID-19 patients and 207 were non-COVID-19 patients. Two groups of patients have significance in comparing age, contact history, leukocyte count, lymphocyte count, C-reactive protein, erythrocyte sedimentation rate (p<0.10). Multiple logistic regression analysis showed age, contact history and decreasing lymphocyte count could be used as individual factor that has diagnostic value (p<0.05). The AUC of first RT-PCR test was 0.84 (95% CI 0.73-0.89), AUC of cumulative two times of RT-PCR tests was 0.92 (95% CI 0.88-0.96) and 0.96 (95% CI 0.93-0.99) for cumulative three times of RT-PCR tests. Ninety-six patients showed typical pneumonia radiological features in first CT scan, AUC was 0.74 (95% CI 0.60-0.73). The AUC of patients’ age, contact history with confirmed people and the decreased lymphocytes were 0.66 (95% CI 0.60-0.73), 0.67 (95% CI 0.61-0.73), 0.62 (95% CI 0.56-0.69), respectively. Taking chest CT scan diagnosis together with patients age and decreasing lymphocytes, AUC would be 0.86 (95% CI 0.82-0.90). The age threshold to predict COVID-19 was 41.5 years, with a diagnostic sensitivity of 0.70 (95% CI 0.61-0.79) and a specificity of 0.59 (95% CI 0.52-0.66). Positive and negative likelihood ratios were 1.71 and 0.50, respectively. Threshold of lymphocyte count to diagnose COVID-19 was 1.53×10^9^/L, with a diagnostic sensitivity of 0.82 (95% CI 0.73-0.88) and a specificity of 0.50 (95% CI 0.43-0.57). Positive and negative likelihood ratios were 1.64 and 0.37, respectively.

**Conclusion:** Single RT-PCR test has relatively high false negative rate. When first RT-PCR test show negative result in suspected patients, the chest CT scan, contact history, age and lymphocyte count should be used combinedly to assess the possibility of SARS-CoV-2 infection.

## Introduction

In December 2019, a novel coronavirus-infected pneumonia epidemic outbroke in Wuhan, Hubei province, China. It has spread to all over China and many other countries within a short period of time. Gene sequencing demonstrated this novel virus has 85% homology with the bat severe acute respiratory syndrome coronavirus (bat-SL-CoVZC45)(2-3). World Health organization (WHO) named this novel coronavirus as severe acute respiratory syndrome coronavirus 2 (SARS-CoV-2) and named the disease caused by it as coronavirus disease 2019 (COVID-19)(4). Over 1,000,000 confirmed cases and more than 50 000 death have been reported by Apr 2^nd^, 2020, from more than 100 countries locate in six continents. WHO has already declared the pandemic of COVID-19(5).

SARS-CoV-2 belongs to the β-coronavirus genus. All populations are vulnerable to SARS-CoV-2, however, elder people are more possible to develop critical illness(6). Infection mainly cause lower respiratory tract infection, induce fever, cough, fatigue and shortness of breath. Some patients may develop symptoms like nasal obstruction or running nose, sore throat and diarrhea. Few patients are asymptomatic with unknown reason yet. Critical ill patients would have dyspnea, hypoxemia, multi organ failure and even die because of these(7-9).

Several studies had revealed epidemiological, radiological and laboratory exmaination characteristics of COVID-19, provided us with some basic understanding of this new disease(10-12). However, most of them are descriptive studies and there was no analysis related to the association of early stage radiological examination and early laboratory tests to the diagnosis. They may contribute to an early diagnostic strategy. At present, real-time reverse transcriptase polymerase chain reaction (RT-PCR) is used as main method of virus infection detection(13). Many studies had shown that the positive rate of RT-PCR could be low, especially the first time of nucleic acid test(12, 14). How to increase the effectiveness of COVID-19 early diagnosis when RT-PCR has high false negative rate is the major challenge during the combat against SARS-CoV-2 around world.

## 1 Materials and Methods

### 1.1 Study type

Bidirectional cohort study.

### 1.2 Objects

All suspected patients that were hospitalized in Xiangyang No.1 Poeple’s Hospital until Feb 9^th^, 2020, follow up was until Mar 20^th^, 2020. COVID-19 patients were confirmed according to the *Diagnosis Guidance for Novel Coronavirus Pneumonia the 6*^*th*^ *Edition* published by National Health Commission of the People’s Republic of China and National Administration of Traditional Chinese Medicine. Suspected patients will have repeat RT-PCR tests with time interval of 24 hours at least. Patients information and data were followed-up until Mar 20^th^, 2020. The study was approved by the ethics review board at Xiangyang No.1 People’s Hospital (No. 2020GCP012). Conventional informed consent was not necessary in this study, due to the emergency outbreak and bidirectional nature of the study, informed consent was waived. All procedures performed in studies involving human participants were in accordance with the ethical standards of the institutional and national research committee and with the 1964 Helsinki declaration and its later amendments or comparable ethical standards.

### 1.3 Data collection

Data collected as follow: the baseline information of all patients, such as gender, age, contact history, time of symptom onset and main manifestations and so on. Chest CT scan and radiological diagnosis. Hemograms data such as neutrophils count, lymphocytes count, monocytes count and so on. First-time laboratory testing results like creatinase, erythrocyte sedimentation rate (ESR), procalcitonin and so on. The final diagnosis based on RT-PCR, and the needed times of tests to get final positivity.

### 1.4 Statistical analysis

SPSS 22.0 and MedCalc were applyed for statistical analysis. Measurement data were described as mean plus standard deviation. Enumeration data were described as number of cases. Significance of measurement data were tested by using *χ*^*2*^*-test*, and *t-test* for enumeration data, *a* was set as 0.10 and *p<a* would be significant. After, all indexes with statistical significance were analyzed with COVID-19 diagnosis results by multiple logistic regression, *a* was set as 0.05 and *p<a* would be seen as significance exists to evaluate the ability of the index could be used as individual diagnostic factor of COVID-19. Applying the area under curve (AUC) of receiver operating characteristic (ROC) curve and 95% confidence interval (CI) to evaluate the diagnostic value of the single index. Based on the AUC we got, indexes were grouped into no diagnostic ability (AUC ≤0.5), low diagnostic ability (0.5<AUC≤0.7), medium diagnostic ability (0.7<AUC≤0.9) and high diagnostic ability (0.9<AUC≤1.0). The thresholds of continuous variables were calculated by Youden’s J statistic, which is sensitivity plus specificity minus one.

## 2 Results

### 2.1 Epidemiological characteristics of the included study objects

There are 315 suspected cases in total, 108 COVID-19 patients are finally confirmed, and 207 cases are non-COVID-19 patients. In these 108 confirmed cases, there are 53 males and 55 females. The youngest patient is 1 year and 6 months old, the oldest patients is 90 years old, and only 2 patients are aged below 18. The average age of these 108 patients is 50.30 (SD = 17.43) years old, many patients are aged from 50 to 70 years old. There are 41 (38%) cases that have underlying diseases, mainly including hypertension, diabetes, coronary heart disease, tuberculosis, hepatitis B, chronic bronchitis, etc. 17 cases are defined as imported cases from Wuhan, 12 cases have travel history to Wuhan, 53 cases have a history contact with the people returned from Wuhan, 30 cases are from family clusters, 24 cases don’t have clear contact history, 2 cases not known. In the 72 cases that have clear contact history, the shortest incubation period is 1 day, the longest incubation period is 20 days. Based on maximum likelihood approach, the incubation period can be best fitted on a Weibull distribution with an estimated mean of 6.16 (95%CI 5.4-7.4) days and SD of 5.04 (95%CI 4.8-5.6) days). In these 207 non-COVID-19 cases, there are 91 males and 116 females, the youngest is 2 months old, the oldest is 91 years old, the average age is 38.84 (SD = 20.05) years old. There are 86 cases that have travel history in the epidemic area or have contact history with people from the epidemic area.

### 2.2 Confirmed case’s RT-PCR test and first chest CT results

First nucleic acid test revealed 73 test positive cases, cumulative two times of nucleic acid test revealed 90 test positive cases, cumulative three times of nucleic acid test revealed 99 test positive cases. In the remaining 9 cases, 6 cases were tested positive in their fourth nucleic acid test, 3 cases were tested positive in their fifth nucleic acid test. All the confirmed cases have received chest CT examination, 96 cases have relatively obvious imaging features of COVID-19.

### 2.3 Characteristic difference bewteen COVID-19 and non-COVID-19 cases

See table 1 of epidemiological characteristics, blood routine examination results before hospitalization, first enzyme level test results after hospitalization of these two groups. Results show that there are statistically significant differences (*P < 0*.*1*) in age, contact history, Creatine kinase (CK), White blood cell (WBC), Neutrophil (Neu), Lymphocyte (Lym), C-reactive protein (CRP) and Erythrocyte sedimentation rate(ESR) between these two groups.

**Table 1.**
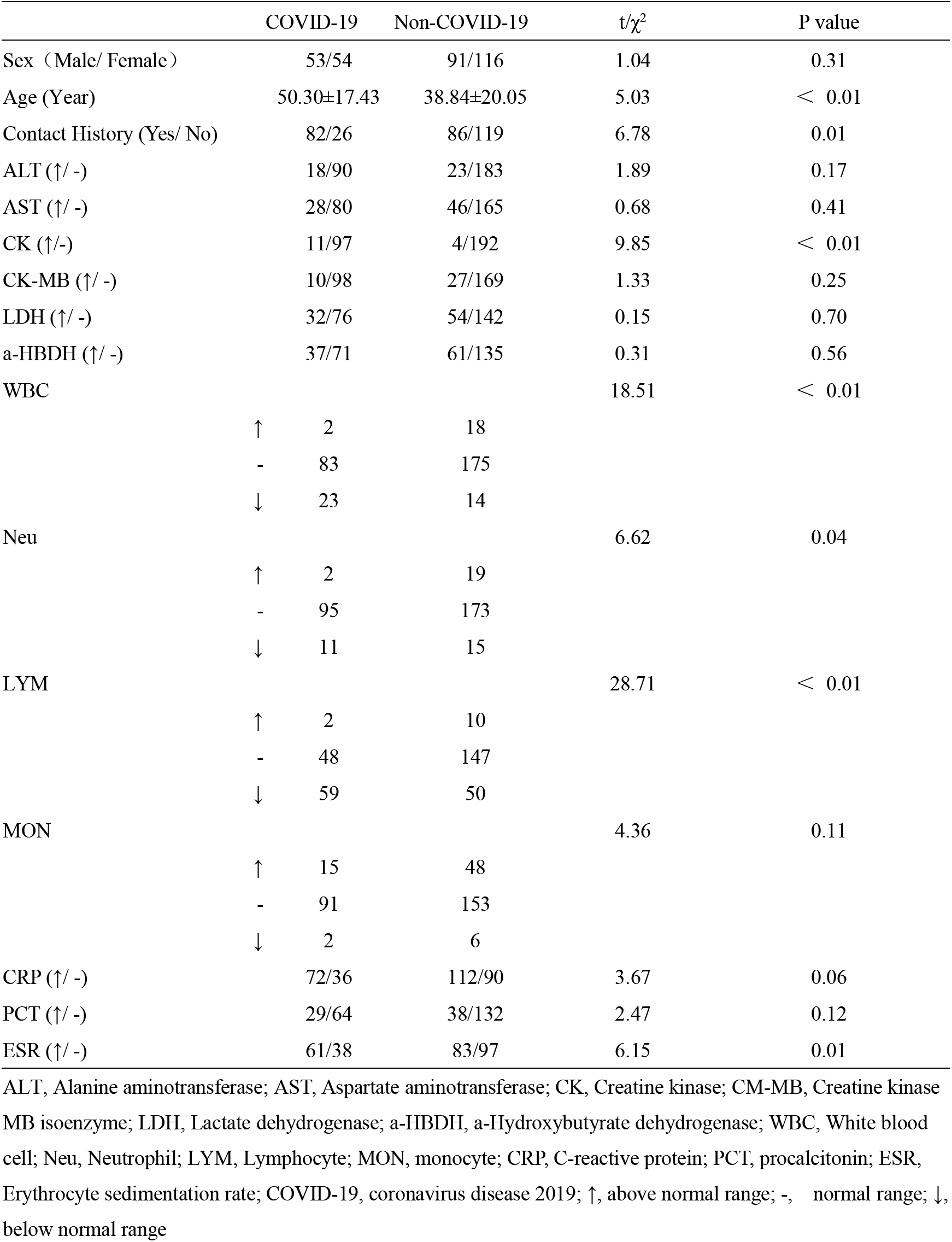
Baseline characteristics and single factor analysis of COVID-19 patients.

### 2.4 Multiple logistic regression analysis

Based on multiple logistic regression analysis, independently age, contact history, and decrease in lymphocyte count have value for COVID-19 diagnosis (Table 2).

**Table 2.** Results of multiple logistic regression analysis.

|  | *B* value | *SD* | *Wals* value | *P* value | *OR* | 95% *CI* |  |
| --- | --- | --- | --- | --- | --- | --- | --- |
|  |  |  |  |  |  | Lower limit | Upper limit |
| Age | 0.034 | 0.009 | 13.247 | 0 | 1.034 | 1.016 | 1.053 |
| Contact History | 1.361 | 0.308 | 19.495 | 0 | 3.901 | 2.132 | 7.138 |
| CK | 0.234 | 0.257 | 0.832 | 0.362 | 1.264 | 0.764 | 2.092 |
| WBC | — | — | 3.831 | 0.147 | — | — | — |
| WBC (1) | -1.214 | 1.55 | 0.613 | 0.433 | 0.297 | 0.014 | 6.198 |
| WBC (2) | -0.134 | 1.659 | 0.007 | 0.936 | 0.875 | 0.034 | 22.597 |
| Neu | — | — | 4.706 | 0.095 | — | — | — |
| Neu (1) | 2.367 | 1.227 | 3.719 | 0.054 | 10.667 | 0.962 | 118.253 |
| Neu (2) | 1.661 | 1.424 | 1.362 | 0.243 | 5.265 | 0.323 | 85.73 |
| LYM | — | — | 7.978 | 0.019 | — | — | — |
| LYM (1) | 0.832 | 1.641 | 0.257 | 0.612 | 2.298 | 0.092 | 57.305 |
| LYM (2) | 1.723 | 1.666 | 1.07 | 0.301 | 5.603 | 0.214 | 146.761 |
| CRP | -0.016 | 0.336 | 0.002 | 0.963 | 0.984 | 0.51 | 1.901 |
| ESR | 0.219 | 0.32 | 0.467 | 0.494 | 1.245 | 0.664 | 2.333 |
| Constant value | -5.382 | 1.466 | 13.48 | 0 | 0.005 | — | — |

CK, Creatine kinase; WBC, White blood cell; Neu, Neutrophil; LYM, Lymphocyte; MON, monocyte; CRP, C-reactive protein; SD, standard deviation; OR, odds ratio; CI, confidence interval; —, No data

### 2.5 ROC curve analysis of indicators with independent diagnostic value

Based on multiple logistic regression results and further evaluation of first three times RT-PCR test results together with chest CT examination results, ROC curves are built to evaluate the above -mentioned indicator’s diagnostic value of COVID-19, see table 3. It is shown that age, contact history and decreased lymphocyte count have certain prediction value, but the diagnostic efficiency is rather low (0.5 < AUC ≤ 0.7). While chest CT examination result and first time RT-PCR test result have medium diagnostic value (0.7 < AUC ≤ 0.9), three and more than three times of RT-PCR test have relatively high diagnostic value (AUC=0.96, 95% CI 0.93-0.99). Chest CT examination result together age, contact history and decrease in lymphocyte count have AUC of 0.84 (95%CI 0.72-0.90). See figure 1 and figure 2.

**Table 3.** The area under curve of diagnostic indexes and combined multiple indexes of COVID-19 diagnosis.

| Diagnostic variability | AUC | SE | 95% CI |  |
| --- | --- | --- | --- | --- |
|  |  |  | Lower limit | Upper limit |
| Age | 0.664 | 0.032 | 0.602 | 0.726 |
| Contact History | 0.637 | 0.033 | 0.573 | 0.702 |
| Hypolymphemia | 0.624 | 0.033 | 0.558 | 0.69 |
| RT-PCR 1 | 0.838 | 0.028 | 0.782 | 0.893 |
| RT-PCR 2 | 0.917 | 0.022 | 0.875 | 0.959 |
| RT-PCR 3 | 0.958 | 0.016 | 0.928 | 0.989 |
| CT | 0.742 | 0.028 | 0.687 | 0.797 |
| NEW | 0.847 | 0.021 | 0.804 | 0.889 |

RT-PCR 1, The first RT-PCR test; RT-PCR 2, The cumulative twice RT-PCR test; RT-PCR 3, The cumulative three times RT-PCR test; CT, computed tomography; NEW= First chest CT scan + Age + Contact History + Hypolymphemia; AUC, Area under subject operating curve; SE, standard error, CI, confidence interval

**Figure 1.**
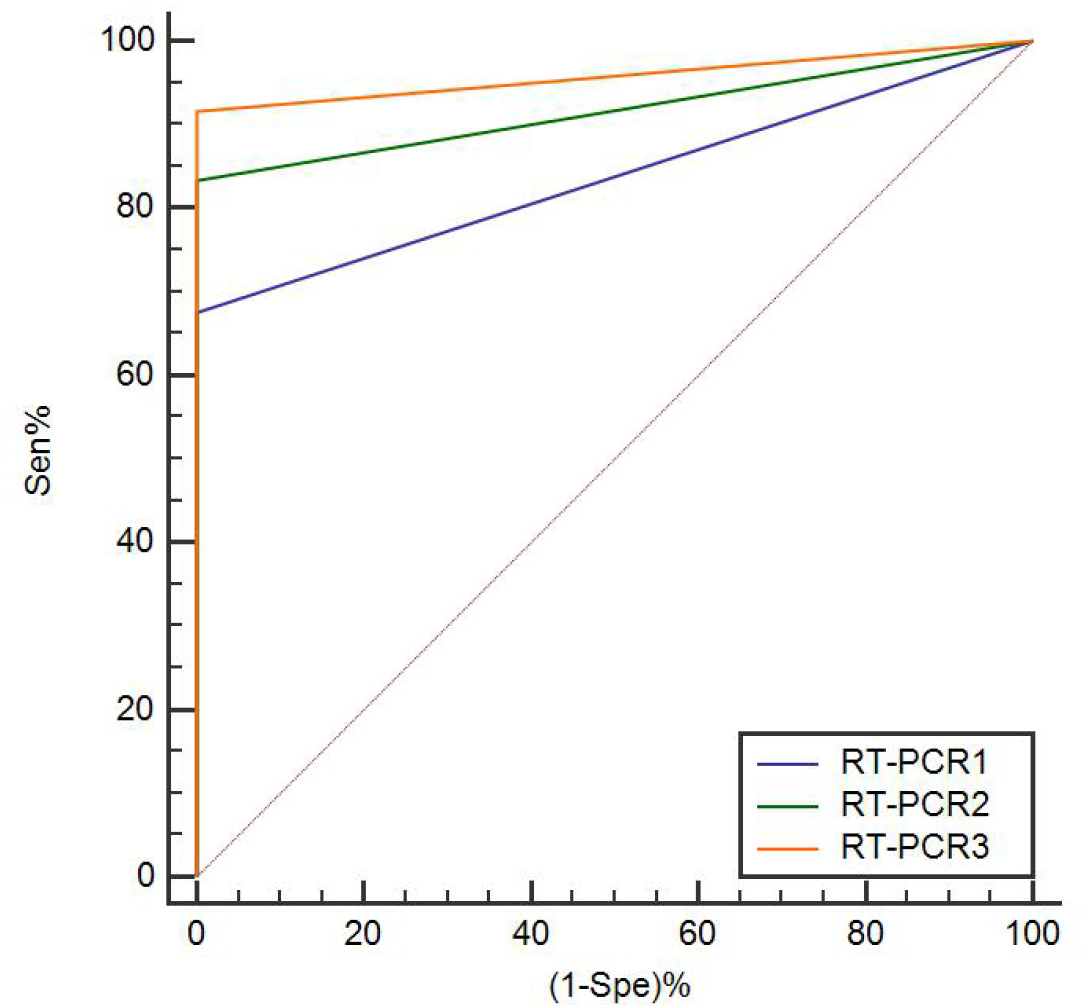
The receiver operating characteristic curve of cumulative three times of RT-PCR tests RT-PCR 1, The first RT-PCR test; RT-PCR 2,The cumulative twice RT-PCR test; RT-PCR 3, The cumulative three times RT-PCR test

**Figure 2.**
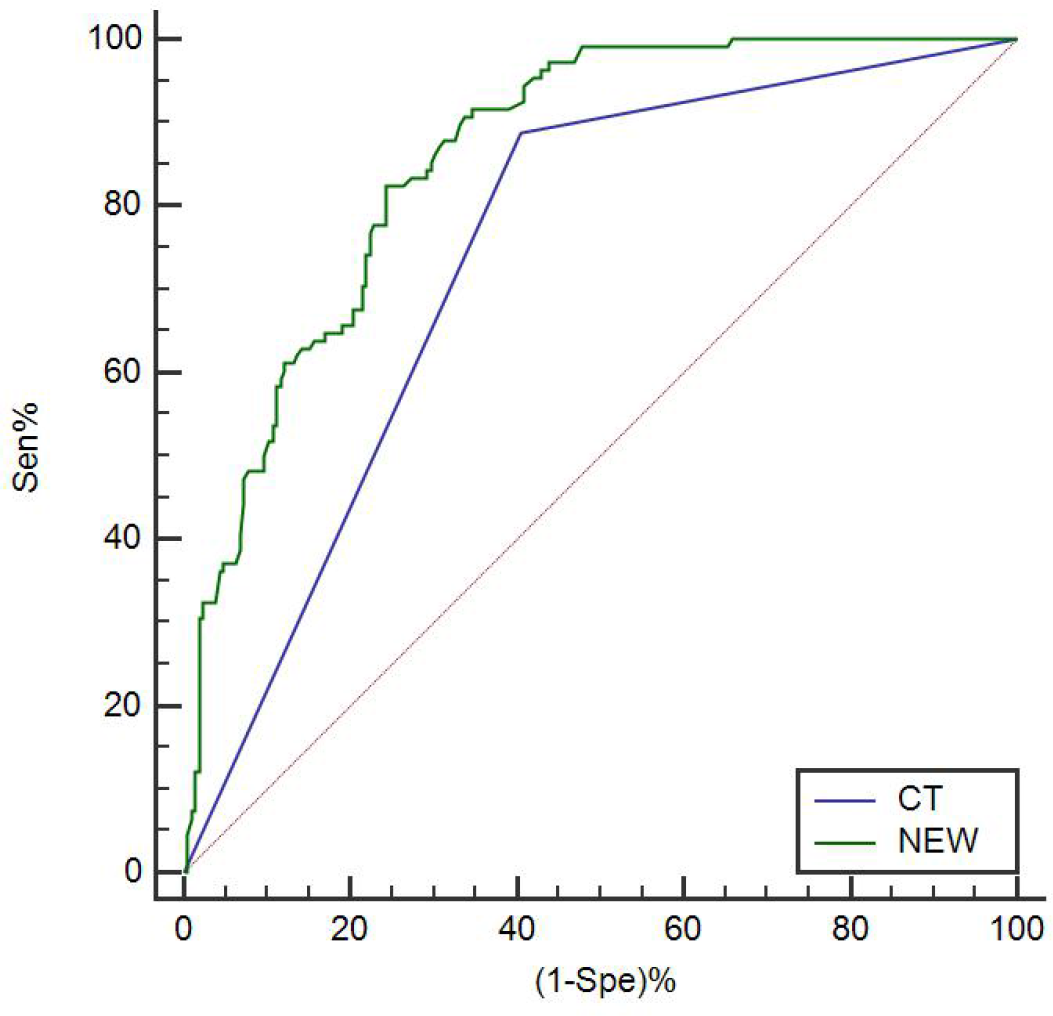
The receiver operating characteristic curve of first chest CT scan and the combination of patients age, contact history and lymphocytopenia NEW= Fist chest CT scan + Age + Contact History + Hypolymphemia; Sen, sensitivity; Spe, specificity

### 2.6 Threshold diagnostic value of patient age and lymphocyte count

In these indicators with independent diagnostic value of COVID-19, age and lymphocyte count are measurement data, threshold analysis showed 41.5 year-old age is the prediction threshold value of COVID-19, with diagnostic sensitivity of 0.70 (95%CI 0.61-0.79), specificity of 0.59 (95%CI 0.52-0.66), positive likelihood ratio of 1.71 and negative likelihood ratio of 0.50. Lymphocyte count of 1.53×10^9^/L is the prediction threshold value of COVID-19, with diagnostic sensitivity of 0.82 (95%CI 0.73-0.88), specificity of 0.50 (95%CI 0.43-0.57), positive likelihood ratio of 1.64 and negative likelihood ratio of 0.37.

## 3 Discussion

SARS-CoV-2 is a new beta-type coronavirus that mainly causes lower respiratory tract infections, specificity of clinical characteristics of COVID-19 is not strong, current diagnosis mainly relies on nucleic acid test. However, the positive rate of first test is low, which means there is high false negative rate, research(12, 14) shows the positive rate of first nucleic acid RT-PCR test of COVID-19 patients is only around 30-50%. Repeated tests are often conducted in clinical work to increase the positive rate, but in-depth study on diagnostic accuracy have not been carried out. A recent research(15) shows the median time from COVID-19 patient’s symptom onset to nucleic acid RT-PCR test positive is 20 days, the shortest time is 8 days, and the longest time is 20 days. It is a disadvantage for early diagnosis of this disease when there is a relatively high false negative rate of nucleic acid RT-PCR test and there is also a relatively long time period from symptom onset to test positive. Especially when human-to-human transmission has been clearly acknowledged, such disadvantage makes it difficult for disease control(9). Finding a method that has diagnostic value in the early stage, and conduct early diagnosis, early isolation and early treatment on suspected cases, would not only be beneficial for patient’s recovery, but also can play a vital role in disease control.

This study includes all the suspected COVID-19 cases that were admitted to our hospital before February 9, 2020, follow up was until March 20. Based on final nucleic acid RT-PCR test results as diagnosis criteria, 108 COVID-19 cases were finally confirmed, and 207 cases were non-COVID-19 cases. The epidemiological characteristics, chest CT examination result and blood routine test result before hospitalization, and first routine laboratory test result after hospitalization can reflect an early status of the COVID-19 patients on a certain degree. Comparing the differences between COVID-19 cases and non-COVID-19 cases in the aspects mentioned above, and further finding indicators with independent diagnostic value through logistic regression analysis would be beneficial for early diagnosis of COVID-19. Especially when there is false negative in the nucleic acid RT-PCR test, these indicators would present their prediction value. Therefore, this study has important practical significance.

In the 108 confirmed COVID-19 cases of this study, 73 of them had their first nucleic acid tested positive, 35 of them had false negative result. First nucleic acid RT-PCR test only has medium diagnostic value (AUC = 0.838), with positive rate of 68% and false negative rate of 32% (35/108), diagnostic sensitivity is 0.68 (73/108). Cumulative two times of nucleic acid RT-PCR test revealed 90 positive cases, which has a relatively high diagnostic efficiency (AUC=0.917), the diagnostic sensitivity is 0.83 (90/108), but there left 18 false negative cases. Cumulative three times of nucleic acid RT-PCR test can reach a relatively ideal diagnosis efficiency (AUC = 0.958), the diagnostic sensitivity is 0.92 (99/108). Therefore, this study also confirms the relatively high false negative rate, which can be miss diagnosis rate, of first nucleic acid test. Based on that, it takes several repeated tests to reach a relatively high diagnosis accuracy. In clinical practice, if suspected cases do not have tested positive, at least three times of nucleic acid RT-PCR test should be conducted for them, in order to decrease miss diagnosis rate.

There are 96 cases that have relatively obvious COVID-19 imaging features in their first chest CT examination result, which indicates chest CT diagnostic sensitivity is relatively high, which is 0.89 (96/108). With AUC = 0.724 also shows that independent chest CT diagnosis has medium diagnostic value. Multiple logistic regression analysis result shows that patient’s age, contact history and decrease in lymphocyte count have independent diagnostic value of COVID-19. Although these indicator’s diagnosis efficiency is relatively low (0.5 < AUC ≤ 0.7), combining chest CT result with age, contact history and decrease in lymphocyte can have a similar diagnosis accuracy with first nucleic acid test (AUC = 0.859). Therefore, these indicators are of great reference importance when nucleic acid RT-PCR test provides false negative result, or before the disease can be detected by nucleic acid RT-PCR test. By combing chest CT result with age, contact history and decrease in lymphocyte to diagnose COVID-19, miss diagnosis rate can be significantly reduced. In clinical work, when diagnosing patients with respiratory symptoms such as fever, cough, and sputum, the patient’s chest CT imaging features should be paid attention to. Moreover, patient’s age should be fully concerned; patient’s anamnesis and epidemiological history (travel history in epidemic area, contact history with people from epidemic area or with confirmed cases) should be carefully inquired; and patient’s lymphocyte count of blood routine test should be taken into consideration as well. This study further finds that age older than 41.5 years old and lymphocyte count lower than 1.53×10^9^/L are the threshold diagnostic numbers for age and lymphocyte count respectively. They have relatively high diagnostic values, with sensitivity of 0.70 and 0.82 respectively.

Previous research(14) shows the main reasons of nucleic acid RT-PCR test’s false negative result are: 1) Lack of standardized operating procedures for specimen collection, storage and processing in clinical work; 2) Different viral loads at different disease stages of COVID-19 have certain effects on nucleic acid RT-PCR test; 3) Lack of calibration test for current nucleic acid RT-PCR test; 4) SARS-CoV-2 may have relatively high mutation rate. Nucleic acid RT-PCR test is the diagnosis basis of confirming COVID-19, but the first-time test has relatively high false negative rate. Diagnosing suspected cases before they are test confirmed can only rely on patient’s clinical characteristics, chest CT imaging features and laboratory test results. Therefore, this study’s conclusion provides a reliable theoretical basis for the clinical diagnosis of COVID-19. Respiratory infection symptoms such as fever and cough, typical chest CT imaging features, middle aged and elderly patients (age > 41.5 years), lymphocyte count lower than 1.53×10^9^/L should be considered linked to COVID-19. If there is available travel history in epidemic area or contact history with people form epidemic area or with confirmed patients, that could provide another strong evidence for diagnosing COVID-19. For these patients, isolation and medical treatment should be carried out as early as possible, which would be helpful for better control of the infection and cut off the transmission at an early stage. Moreover, this study’s first-time nucleic acid RT-PCR tests have a test positive rate of 68%, which is higher than previous study(14, 16) (positive rate 30-50%). It may be related to the fact that the nucleic acid test samples were collected and stored by specialized personnel in our hospital who had received professional training. Which indicates that nucleic acid test in strict accordance with operating procedures, careful sample storage and transfer procedures can increase the positive rate of first-time nucleic acid test, therefore the miss diagnosis rate of COVID-19 can be reduced and the confirmation time can be shortened. This is also of great importance for early diagnosis of the disease and epidemic control.

This study provides new theoretical basis for diagnosis on COVID-19, especially under the circumstance that there is relatively high false negative rate of nucleic acid RT-PCR test, and the time period from symptom onset to disease test detectable can be long, this study’s result is of great reference significance. This study’s result is helpful for early diagnosis, early isolation and early treatment of the patients, and can also play an important role in better epidemic control. However, this study has certain limitations. Firstly, this study’s sample size is limited, which may lead to some bias in the research result. Secondly, this study only studies on patient’s chest CT results before hospitalization, blood routine test results and first routine laboratory test results after hospitalization. Other indicators that may have diagnostic value for early diagnosis might be missed. Thirdly, previous research(15) shows few patients can have longest time period of 38 days from symptom onset to nucleic acid tested positive. While in this study the follow up lasted for 30 days, therefore it is possible of miss diagnosis in the cases that are included in this study. However, this study is a two-way cohort study, from the aspects of etiology and diagnostic accuracy test, the study result is still reliable. Moreover, this study mainly focuses on patient’s epidemiological characteristics, chest CT result and blood routine test result before hospitalization, and first routine laboratory test result after hospitalization, which can reflect the early status of patients after symptom onset more accurately. And it is of high reference value for early diagnosis. Currently, previously established cohort is still under further follow up, which will provide more detailed information for early diagnosis, disease characteristics and prognosis, in order to have a more in-depth understanding of COVID-19.

### 4 Conclusion

Single RT-PCR test has relatively high false negative diagnostic rate. Cumulative three times of nucleic acid test would give high positive rate of virus detection. When first RT-PCR test show negative result, the chest CT scan, contact history, age and lymphocyte count of the suspected patient should be used combinedly to assess the possibility of SARS-CoV-2 infection.

## Data Availability

All data in the manuscrpt are availability

## Abbreviations

COVID-19: novel coronavirus disease 2019
SARS-CoV-2: severe acute respiratory syndrome coronavirus 2
RT-PCR: real time reverse transcriptase polymerase chain reaction
ROC: receiver operating characteristic
AUC: area under the receiver operating characteristic curve
WHO: World Health organization
ALT: Alanine aminotransferase
AST: Aspartate aminotransferase
CK: Creatine kinase
CM-MB: Creatine kinase, MB isoenzyme
LDH: Lactate dehydrogenase
a-HBDH: a-Hydroxybutyrate dehydrogenase
WBC: White blood cell
Neu: Neutrophil
LYM: Lymphocyte
MON: monocyte
CRP: C-reactive protein
PCT: procalcitonin
ESR: Erythrocyte sedimentation rate
Sen: sensitivity
Spe: specificity

